# Yellow fever outbreak potential in Djibouti, Somalia and Yemen

**DOI:** 10.1101/2024.08.07.24311590

**Authors:** Keith Fraser, Laurence Cibrelus, Jennifer Horton, Chiori Kodama, J. Erin Staples, Katy A. M. Gaythorpe

**Author notes:** Corresponding authors and; Department of Infectious Disease Epidemiology, Imperial College London, 90 Wood Lane, London W12 0BZ, UK.

## Abstract

The importation of arbovirus diseases into countries where they are not currently endemic is a global concern, driven by human movement and direct and indirect climate change effects. In the World Health Organization Eastern Mediterranean region, three countries - the Republic of Djibouti, the Federal Republic of Somalia, and the Republic of Yemen - are currently considered to be at potential or moderate risk for yellow fever outbreaks, and an assessment for outbreak potential in the event of importation was sought. Djibouti and Somalia share land borders and significant cross-border movement with countries where yellow fever is endemic, while Yemen is currently experiencing a crisis which has severely impacted healthcare infrastructure, and has already seen suspected outbreaks of other similar arboviruses such as dengue, chikungunya and West Nile.

Here we present a mathematical modelling assessment of the risk of introduction and propagation of yellow fever in Djibouti, Somalia and Yemen. This modelling has two components: projecting the risk of importation of infectious individuals into individual administrative regions of the countries of interest, and the use of a dynamic yellow fever model to model yellow fever virus transmission within the same regions.

We present results showing that certain regions of Djibouti, Somalia and Yemen are at higher risk than others for yellow fever outbreaks, with the risk being higher in some areas such as the western coastal regions of Yemen (an area that has experienced recent outbreaks of other arboviruses), regions of Somalia bordering both the Federal Democratic Republic of Ethiopia and the Republic of Kenya, and Djibouti City.

## 1 Introduction

Yellow fever (YF) is a viral haemorrhagic fever endemic in tropical regions of Africa and South America with rapid propagation and amplification, particularly in urban settings, and potential for international dissemination. With multiple transmission cycles, a sylvatic reservoir in non-human primates (NHPs) and no treatment available, eradication is not feasible; control is primarily carried out through early identification of cases, vaccination of susceptible people and vector control activities. Targets for YF control therefore focus on outbreaks, particularly in urban settings. These targets are set by the Eliminate Yellow Fever Epidemics (EYE) Strategy [1] whose focus is i) protecting at-risk populations, ii) preventing international spread, and iii) containing outbreaks rapidly. To achieve these aims, a clear categorisation of risk is crucial in order to tailor public health measures to each level of risk..

The risk of YF can be examined in a range of ways [2]. This may be informed by recent outbreaks, current vaccination coverage or environmental conditions to project, for example, the transmission intensity (measured as a basic reproduction number or force of infection) or environmental suitability to the vector. The EYE strategy risk analysis working group has worked extensively on defining appropriate criteria to assess risk [1]. However, depending on the metric used, this risk assessment can vary; for example transmission intensity estimates may be affected by climate change and estimates of under-immunised populations may be affected by recent vaccination activities. Climate change may lead to the introduction of yellow fever virus (YFV) to regions where it is not currently circulating [3]. This is of concern since the population would have no natural or vaccine-induced immunity, creating the potential for large outbreaks. Similarly, human activities such as changing movement patterns, deforestation or changing agricultural practises can lead to changes in virus circulation and human risk. As a result, the risk of YF is in flux.

Assessing and monitoring risk of YF is critical, particularly in areas bordering existing endemic regions. This is to ensure that interventions and surveillance can be strengthened to capture and respond to YF in new areas and populations. Focusing only on current endemic areas could miss potentially vulnerable populations affected in the future. Mathematical modelling, alongside surveillance and other approaches, is a way of bringing together information on the current situation with knowledge of mechanisms for how that situation may change in future, for example, because of changes in the environment or population.

Here we examine the outbreak potential of YF in the Republic of Djibouti, the Federal Republic of Somalia, and the Republic of Yemen, three countries bordering endemic regions and considered to be at moderate or potential risk of YF outbreaks. [1]. Firstly, we capture the potential spatial spread of an outbreak using estimated values of the subnational force of infection and a radiation model of human movement. Secondly, we use a stochastic, dynamic model of YFV transmission to estimate the risk of an outbreak in the regions of interest and the risk of a single imported case leading to an outbreak. In this way, we examine the risk that YFV could be imported into a district (administrative level 2) and then the potential for that case to propagate an outbreak once there.

## 2 Materials and methods

### 2.1 Overview

We examine two components of risk: 1) the risk of introduction/importation and 2) the risk of a propagated outbreak given introduction. To examine the risk of introduction we couple a simple model of human movement, a radiation model, with the known natural history of YFV infection to give a relative risk that someone might move from an endemic area into a new area whilst infectious. We then use this relative risk to examine the potential of an ensuing outbreak using a mechanistic model of YFV transmission and infection. Through the combination of these approaches, we can highlight areas that may be vulnerable to YFV introduction and ongoing transmission.

Figure 1 shows the workflow of estimates. We begin with estimates of the underlying environmental suitability from Fraser et al. [4]; these estimates are used to *either* seed start locations in endemic countries and calculate the risk of introduction in at-risk regions *or* calculate the underlying outbreak risk in at-risk countries. Finally, the relative underlying outbreak risk is combined with the risk of introduction into at-risk regions to provide a relative outbreak risk.

**Fig 1.**
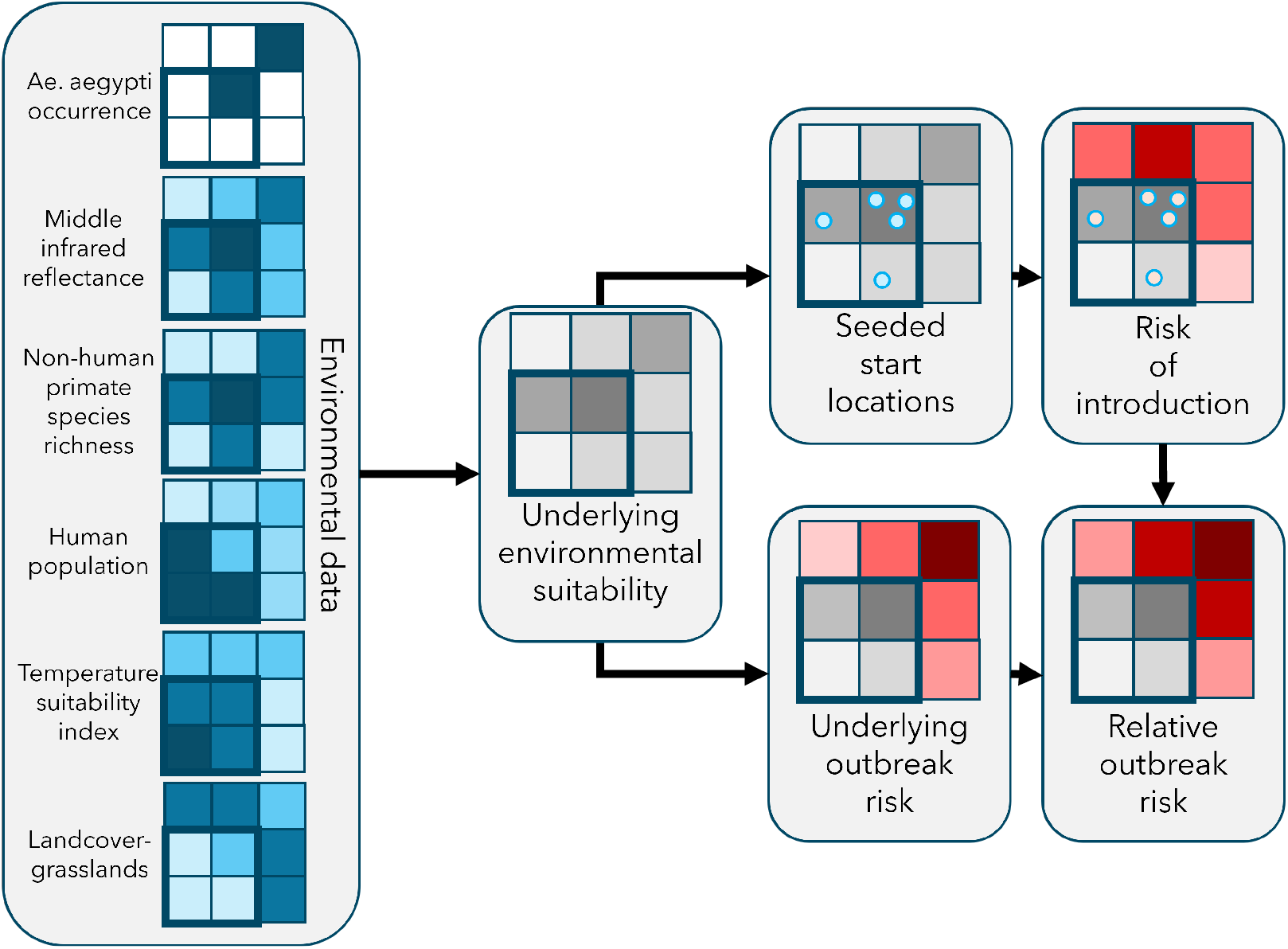
Diagram of methods. Environmental data (left) is used to inform the underlying environmental suitability, defined as transmission intensity (see Fraser et al. for full details [4]). Top: start locations for hypothetical YF outbreaks are seeded proportional to the underlying environmental suitability; the risk of introduction of YFV is estimated for each seeded start location. Bottom: the underlying outbreak risk, defined as the proportion of simulations with a YF outbreak given the underlying environmental suitability; relative outbreak risk, defined as the composition of underlying outbreak risk and risk of introduction. Squares indicate individual districts; darker colour shade implies higher risk (red), higher value of environmental input data (blue), or higher environmental suitability (grey).

### 2.2 Risk of introduction

To examine the potential spatial spread of a YF outbreak we utilise the RAPTORX online application https://shiny.dide.ic.ac.uk/raptorx/. This methodology was initially developed for examining the potential spread of YF in the outbreaks of 2016 in the Republic of Angola and the Democratic Republic of the Congo and has since been further refined. The RAPTORX framework relies on a radiation model of human movement, which depends on the population size and distance between districts [5].

The radiation model performs well at approximating the distribution of long distance movements [6]. This framework can be coupled with information on population immunity (though no pre-existing immunity is assumed in this application) to produce the relative risk of YF occurrence in a particular district, formed from the combined risk of introduction from surrounding districts. In this case, we focus on the risk within districts in Djibouti, Somalia, and surrounding countries. Currently the application focuses on the African continent and so Yemen is not included in importation modelling. This is in part as we assume land-based cross-border movement.

In this analysis we examine the potential spread of an outbreak of YF originating in an unknown region in countries bordering Somalia and Djibouti. We sample from potential start points within the countries directly neighbouring Djibouti and Somalia and propagate the risk of spread to other regions. For each country, we sample fifty start locations, weighted by the force of infection due to sylvatic spillover [4], and propagate the risk of spread from those locations using the radiation model ten times per start point. We then aggregate the results and take the mean (or median in the supplementary material) risk per district across all simulations.

### 2.3 Underlying outbreak risk

We define an outbreak as the occurrence of one or more human YF disease cases with severe symptoms. To model outbreak occurrence, we use a stochastic, dynamic model of YFV transmission that incorporates both sylvatic spillover and human-to-human (urban) transmission [4]. The key parameters for this model are the force of infection for sylvatic spillover, λ_*S*_, and the basic reproduction number of human-to-human transmission, R_0_. These are calculated from environmental covariates (including non-human primate species richness, land cover and temperature range suitability) using parameters estimated within a Bayesian framework from YF seroprevalence and annual reported case data from currently affected regions of Africa and South America.

Figure 2 shows how median values of λ_*S*_ and R_0_ are expected to vary across the three countries considered based on environmental covariate values. 1000 sets of values were drawn from the overall parameter distribution produced from existing YF data [4]; the figures show the median values of λ_*S*_ and R_0_ from the resulting distribution.

**Fig 2.**
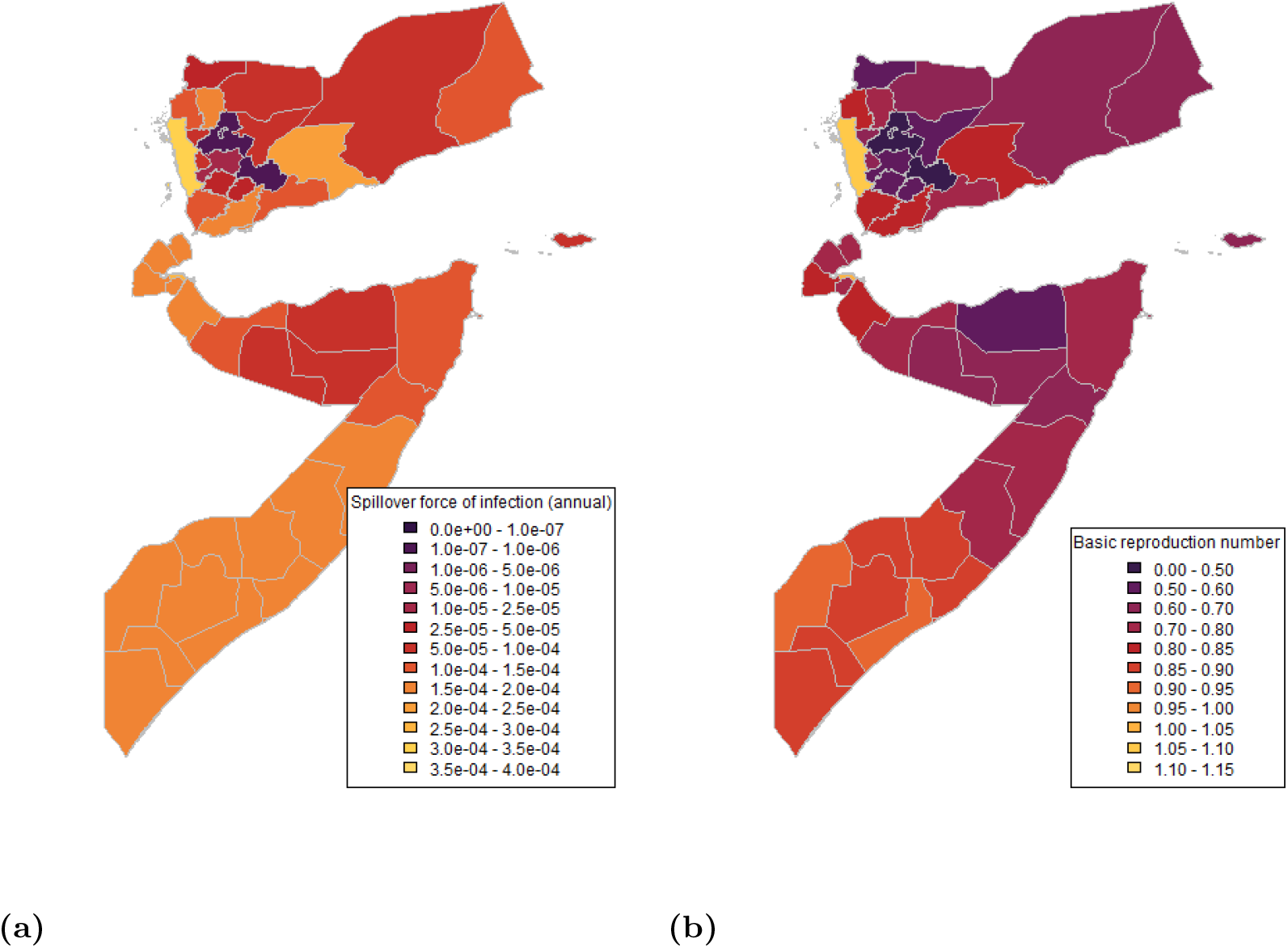
Median values of (a) annual force of infection for sylvatic spillover and (b) basic reproduction number for human-to-human transmission in first-level subnational administrative regions of Djibouti, Somalia and Yemen, calculated from environmental covariates.

We assume the severity spectrum as estimated by Johansson, Vasconcelos and Staples (i.e. that 12% of infections cause severe symptoms [7]). Population sizes are assumed from United Nations World Population Prospects 2019 edition (UNWPP) and distributions are assumed from Landscan 2017 edition [8, 9].

Below we consider a version of underlying outbreak risk where one infectious human is introduced with no sylvatic spillover, based on the assumption that a sylvatic reservoir of YFV is not present in these countries. In the supplementary information we also consider a speculative one in which there is an endemic sylvatic reservoir. The outbreak risk given the calculated epidemiological parameter values is estimated by running the model from the introduction of a single infection with all 1000 sets of R_0_ values (median value shown in Fig. 2a) and recording the proportion of instances in which an outbreak occurs in the target year.

### 2.4 Relative outbreak risk

To calculate the relative risk of an outbreak we multiplicatively combine the risk of potential spread of an outbreak given an infectious individual with the landscape of outbreak occurrence probability. In summary, section 2.2 provides a relative risk of introduction; section 2.3 provides an underlying probability that an introduced infectious individual will propagate an outbreak. The product of these two outputs provides a relative risk of outbreak given introduction.

## 3 Results

### 3.1 Risk of introduction

When we consider the mean risk in Djibouti and Somalia for cases seeded in neighbouring countries (see figures 3 and 6) we find that both countries have relatively low mean risk but have some districts where the simulated risk is non-zero. In Somalia, districts of non-zero risk are generally found in the West, particularly Rab Dhuure in Bakool, which has non-zero risk in over 96% of simulations. In Djibouti, Djibouti City has non-zero risk in over 70% of simulations. See table 1 for the twenty districts that most frequently show non-zero risk.

**Table 1.** The twenty districts with the lowest proportion of simulations with zero risk where cases are seeded in neighbouring countries. Note, this only highlights zero risk and not the magnitude of risk simulated.

| Name | Province | District | Proportion of simulations with zero risk (%) |
| --- | --- | --- | --- |
| Somalia | Bakool | Rab Dhuure | 2.67 |
| Somalia | Gedo | Dolow | 6.67 |
| Somalia | Bari | Qardho | 8.00 |
| Somalia | Bakool | Ceel Barde | 8.67 |
| Somalia | Bay | Qansax Dheere | 8.67 |
| Somalia | Bari | Iskushuban | 11.33 |
| Somalia | Bari | Qandala | 12.00 |
| Somalia | Gedo | Ceel Waaq | 12.00 |
| Somalia | Banaadir | Mogadisho | 15.33 |
| Somalia | Bakool | Tiyeeglow | 16.67 |
| Somalia | Jubbada Dhexe | Jilib | 19.33 |
| Somalia | Gedo | Garbahaaray | 20.67 |
| Somalia | Woqooyi Galbeed | Hargeysa | 20.67 |
| Djibouti | Djibouti | Djibouti | 21.33 |
| Somalia | Gedo | Beled Xaawo | 25.33 |
| Somalia | Bay | Diinsoor | 27.33 |
| Somalia | Awdal | Zeylac | 28.67 |
| Somalia | Bay | Baydhabo | 31.33 |
| Somalia | Gedo | Luuk | 32.00 |
| Somalia | Shabeellaha Hoose | Wanla Weyn | 32.00 |

**Fig 3.**
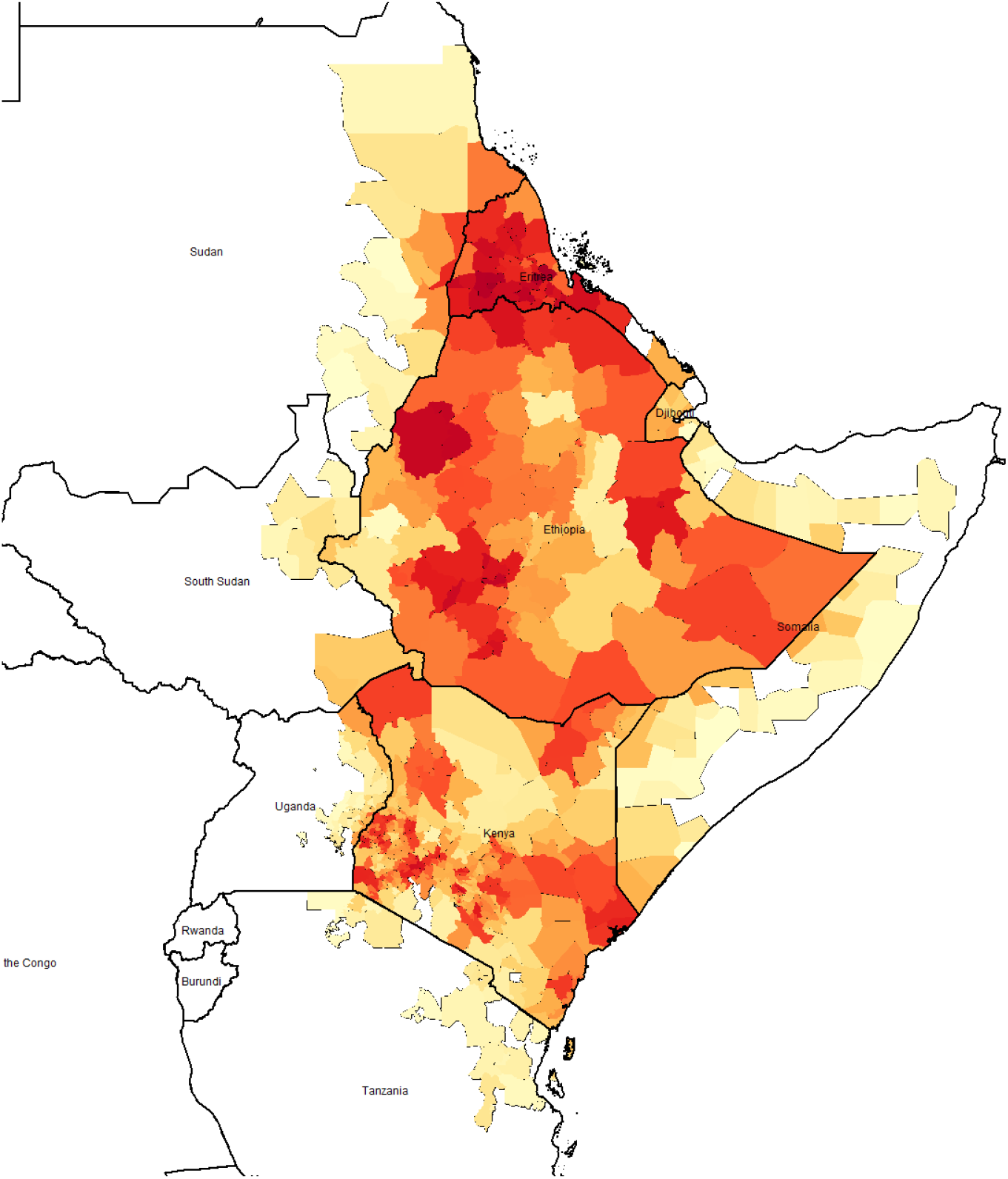
Mean **relative** risk of YFV introduction in second administrative locations given start locations in neighbouring countries where red indicates high risk and white/yellow indicates no/low risk. Median estimates and starting locations are shown in the supplementary material.

If we compare mean and median (supplementary figure 8) risk, there are some notable differences as there are a large proportion of simulations resulting in no estimated risk per district. This highlights the stochastic nature of the resulting potential spread from a starting location.

### 3.2 Underlying outbreak risk

Figure 4 presents the probability of a single case propagating an outbreak. The highest risk (over 80%) is found in coastal western Yemen (Al Hudaydah) and Djibouti City. Other regions with a risk over 50% are found in southern Somalia (Gedo and Lower Shabelle). Figure 9 in the supplementary material shows the risk of an outbreak in 2023 assuming an existing sylvatic reservoir. The outbreak risk is high (*≥*99%) for most regions, except for interior regions of western Yemen.

**Fig 4.**
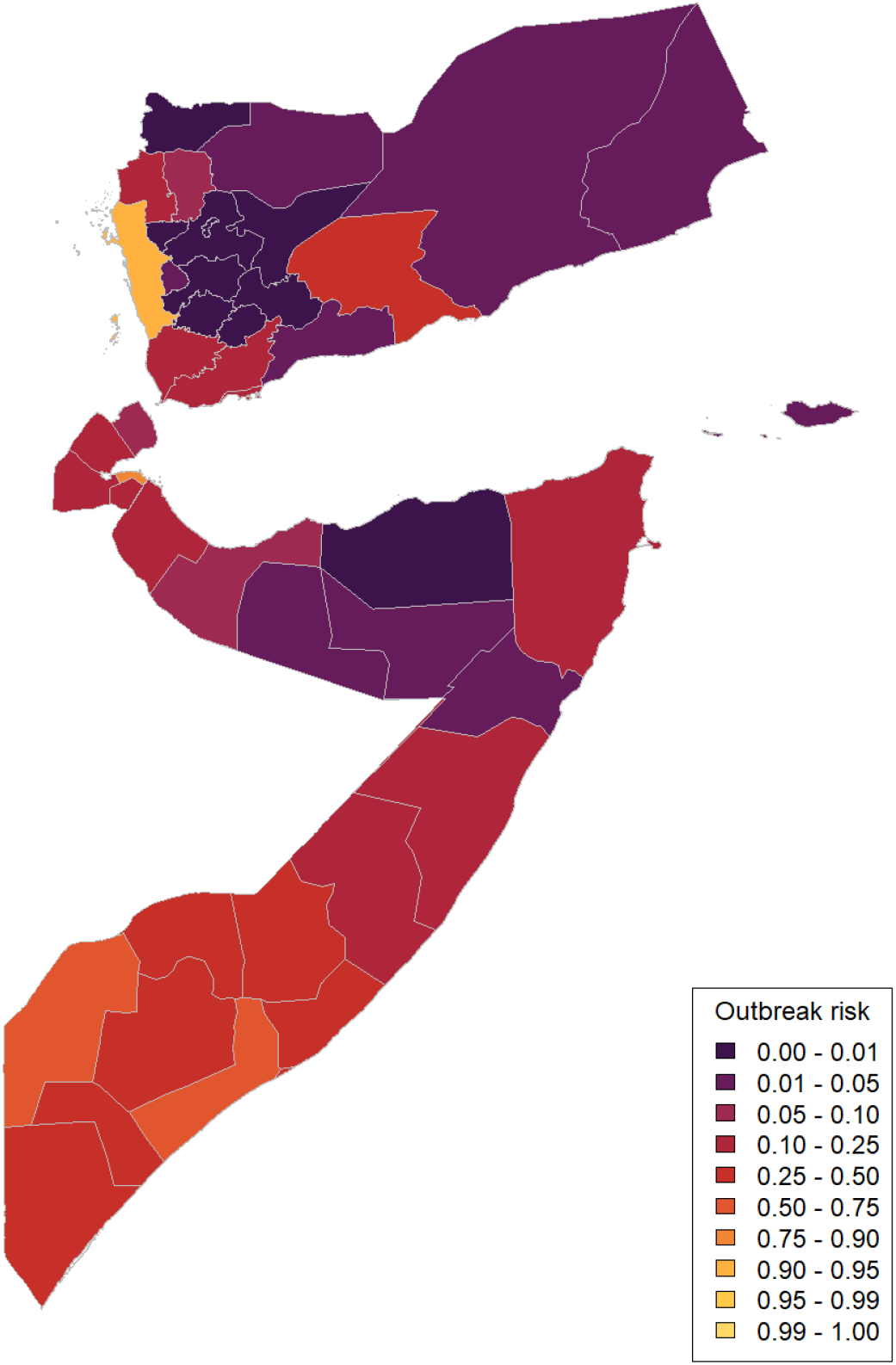
Map of underlying outbreak risk in first level subnational administrative regions in Djibouti, Somalia and Yemen in 2023. This is as calculated as the proportion of simulations where introducing a single infection propagates an outbreak given our underlying estimates of the basic reproduction number from [4].

### 3.3 Relative outbreak risk

Figure 5 presents the final relative risk of an outbreak in Djibouti and Somalia, based on mean introduction risk values. The risk is concentrated in border areas, particularly in the south-west and north-west. An alternative map based on median introduction risk values is shown in 10 in the supplementary information.

**Fig 5.**
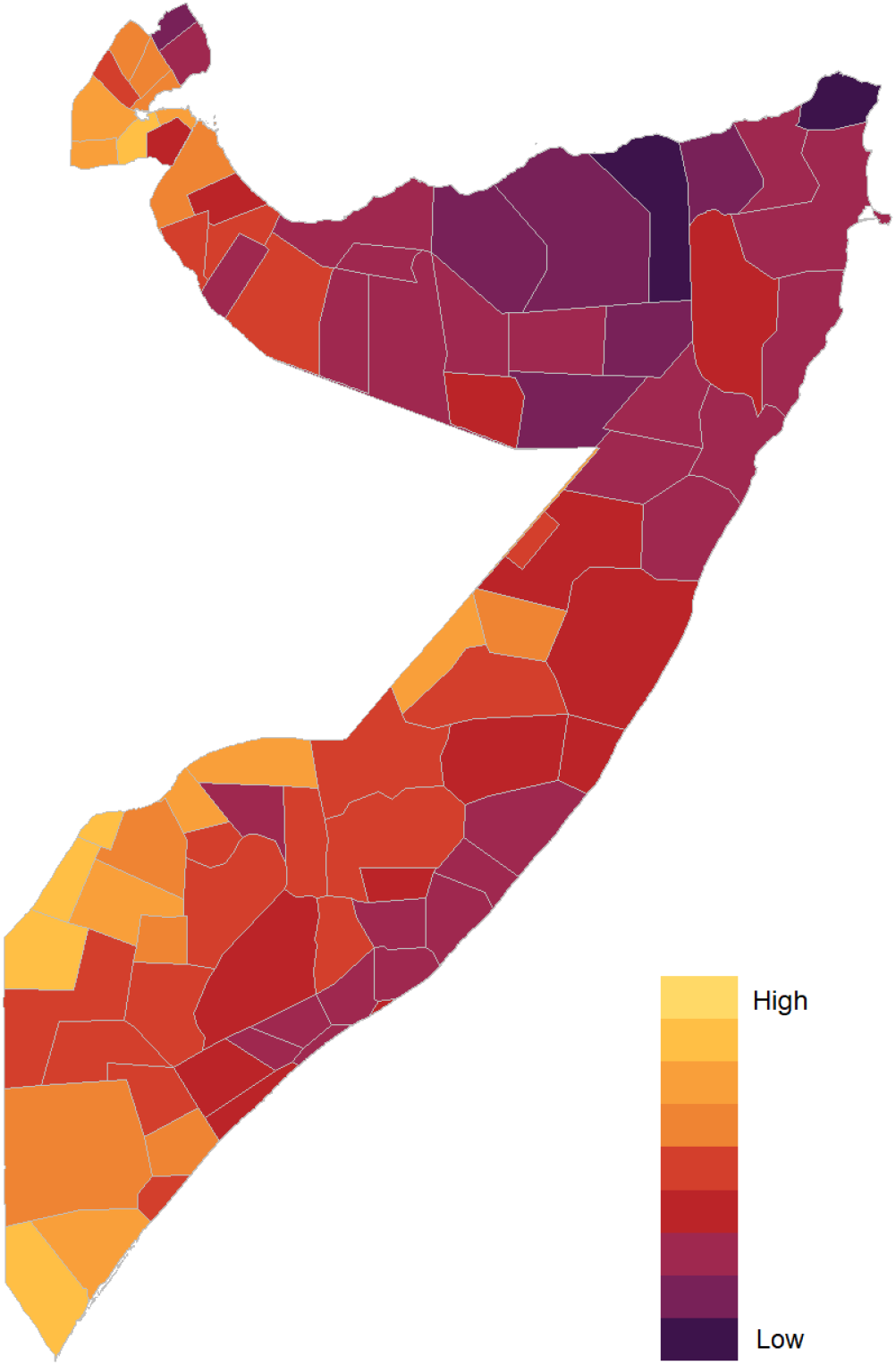
Map of relative outbreak risk in second level subnational administrative regions in Djibouti and Somalia in 2023 calculated via multiplying mean introduction risk scores for each region by the underlying outbreak risk based on a single introduced case for the corresponding first level subnational administrative region

## 4 Discussion

We have examined the risk of spread of an existing YF outbreak to at-risk countries neighbouring or near the endemic zone, as well as examining the outbreak potential of those same at-risk countries. In Djibouti and Somalia there are areas where there is a high likelihood of YFV introduction in the event of an outbreak in neighbouring countries, most notably in Djibouti city and southwest Somalia. When we examine the inherent potential for YFV transmission in Djibouti, Somalia, and Yemen we note the same areas have potential for outbreak propagation but also highlight that a case introduced into Western Yemen has high potential to propagate an outbreak. However, we note the absence of a wildlife reservoir. Our combined model approaches emphasise that there are some districts in Djibouti and Somalia at potential risk of YF outbreaks. This risk is due to geographic proximity and likely transport routes with endemic areas and estimated environmental suitability for YFV transmission. These results emphasise the importance of awareness of potential YF importation to moderate and potential risk countries (as defined by the EYE Strategy [1]) and consideration of YF within testing and diagnosis strategies for compatible clinical presentations.

Dengue is a notable issue in Yemen [10–12], Somalia [13] and Djibouti [14] and shares the same urban vector as YFV. Yet, there has not been a modelled quantification of the risk of YF outbreaks in these countries, such as by the EYE Strategy risk analysis working group. We have highlighted areas in Djibouti and Somalia that have outbreak potential and are vulnerable to YFV introduction, which may have wider implications for surveillance and control activities. Similarly the implications of these results for the prospects of outbreak potential highlight the need for further research to understand vector competence and potential affects of cross-reactive immunity. The region is already affected by other arbovirus outbreaks, as well as disruption of provision of care and dismantling of health systems and services by the ongoing Yemen crisis, and it contains much of Yemen’s population (possibly affected by movement of refugees). Vaccination is the primary form of routine control for YF and the key emergency response in an outbreak, and global stockpiles were expanded in 2016 to hold six million doses for outbreak response [15] (note that this represents only a fraction of the combined populations of the three countries considered). However, if the likelihood of outbreaks in neighbouring regions is altered by human movement and changing environmental factors driven by climate change, stockpiles may need to adjust to be able to address future threats.

There are number of modelling assumptions required to conduct these projections and uncertainty in the input data. The relative risk model of geographic spread assumes that human movement is land-based and motivated only by population sizes and distance between start and destination; however, we do not account for changes over time and by season or alternative travel routes such as air and sea travel. Furthermore, we assume national borders are porous which may, again, vary by region. Regarding the outbreak occurrence model our estimates of the transmission potential based on data from regions with endemic YF. It is worth noting that the existence or endemicity of dengue in these countries, whilst suggestive of a suitable transmission environment, may imply cross-reactivity, which is not accounted for this analysis as we assume populations are fully susceptible. Similarly, whilst we have incorporated some sources of uncertainty, the uncertainty is still underestimated in this analysis, particularly as the countries examined have reported no historic endemic circulation of YFV and thus the projections cannot be validated in these regions.

## 5 Conclusion

Countries and regions bordering existing YF endemic regions are potentially vulnerable to both introduction of YF cases and subsequent outbreak spread. Changing patterns of human movement and displacement, as well as changing environmental conditions through climate change, could exacerbate this risk. We highlight and quantify areas of relatively higher risk in Djibouti, Somalia and Yemen. This complements ongoing work from the WHO EYE Strategy and further emphasises the importance of awareness of YF importation risk when considering testing and diagnosis in compatible clinical presentations.

## Acknowledgments

We would like to acknowledge Arran Hamlet, Giuseppina Ortu, Marie-Eve Raguenaud, and João Paulo Toledo.

## Data availability

Data and R code used in this work is available in the two GitHub repositories below: Risk of geographic spread: https://github.com/mrc-ide/YF_WHO_risk_reports Risk of outbreaks: https://github.com/mrc-ide/DSY_YF_outbreak_risk2

## Supporting information

### Risk of introduction

**Fig 6.**
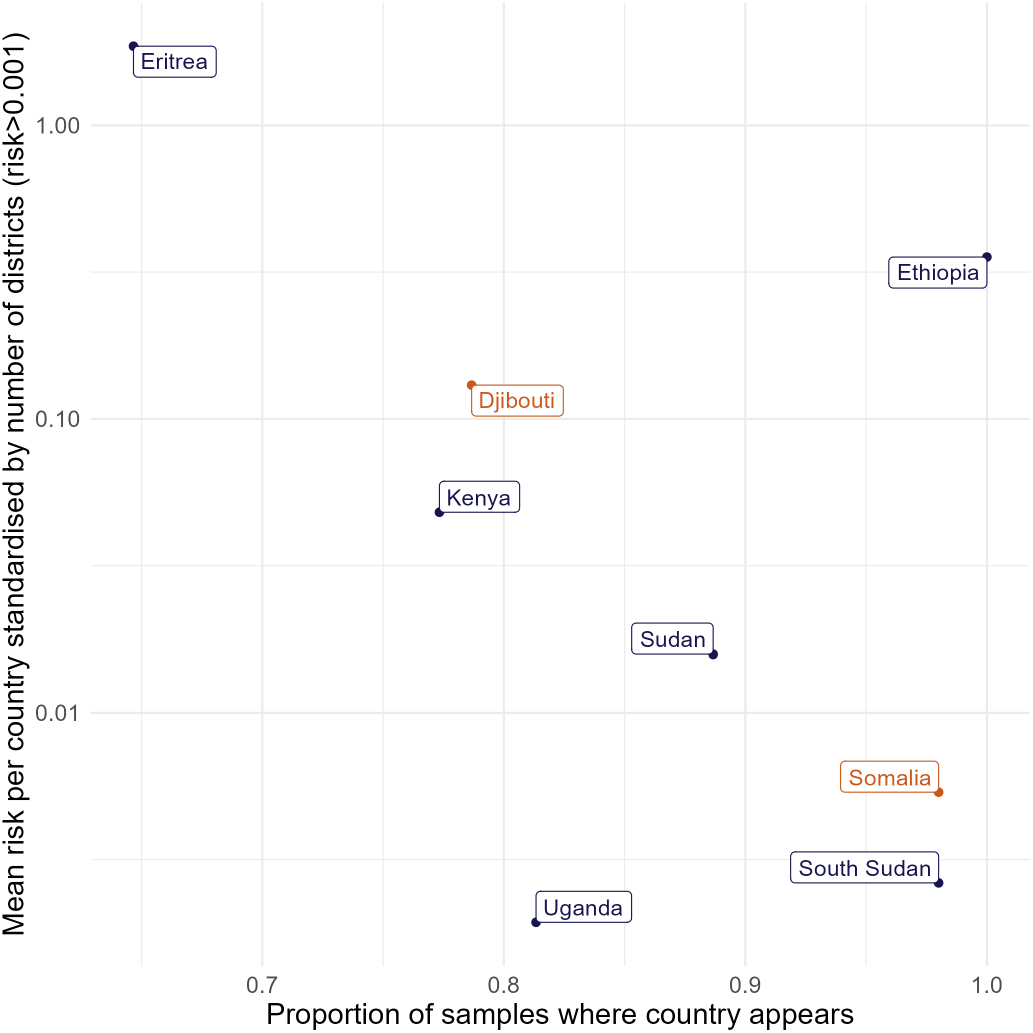
Relationship between the number of times a country appears in simulations and the mean risk score standardised by the number of second administrative locations from start locations in neighbouring countries.

**Fig 7.**
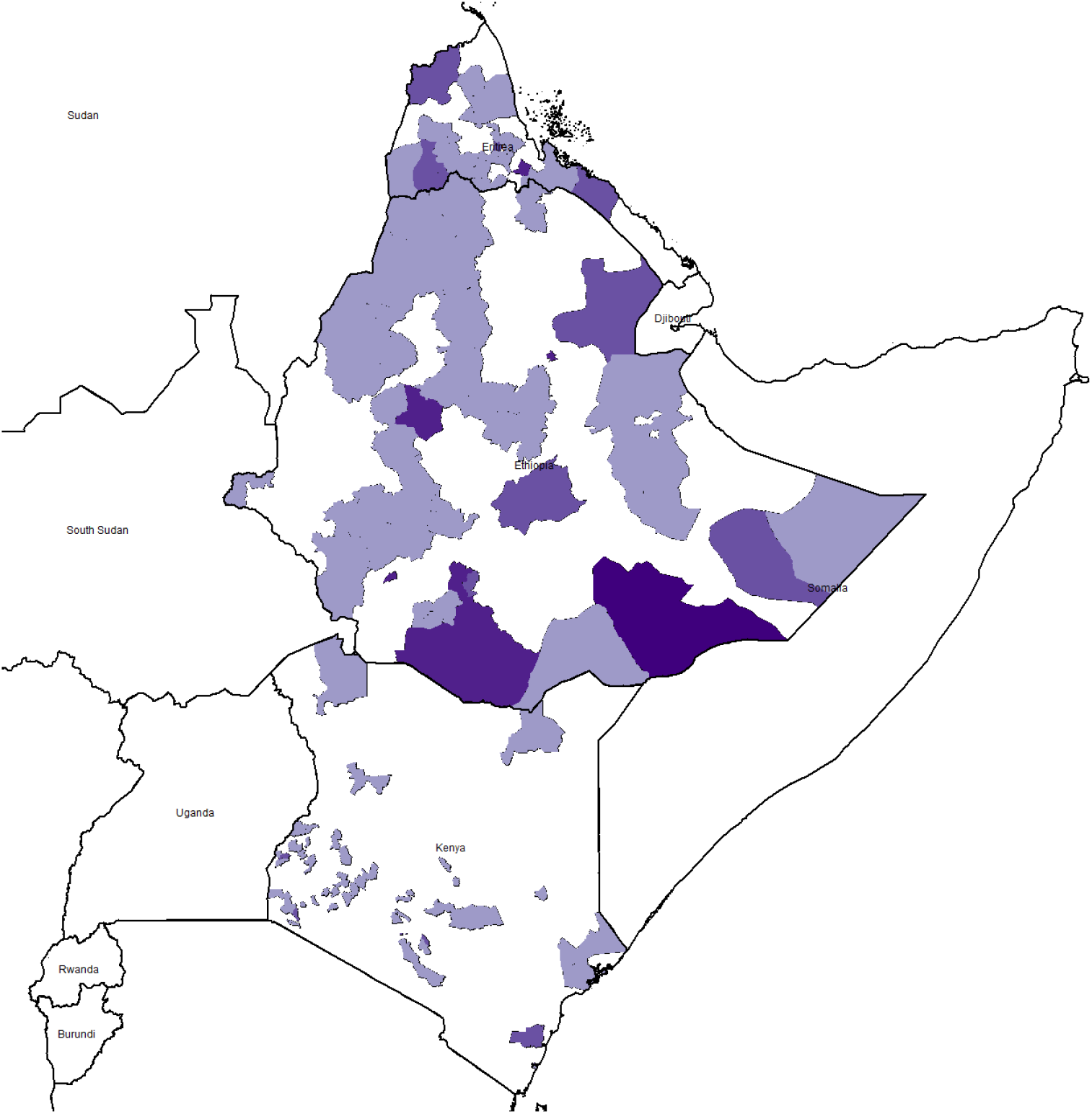
Start locations in neighbouring countries to Djibouti and Somalia. 50 samples were taken per country where a darker colour implies that district was sampled at a higher frequency.

**Fig 8.**
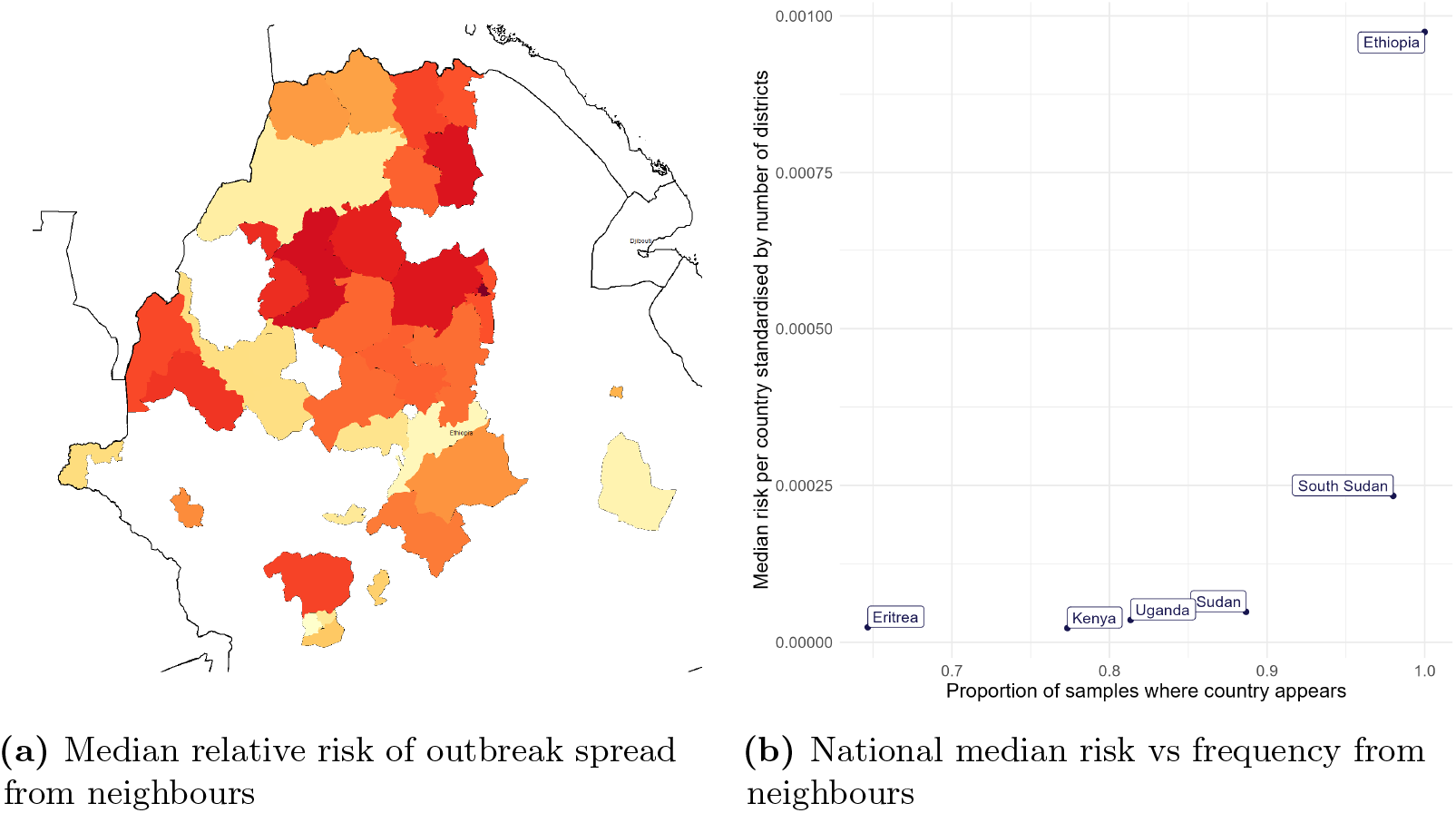
Median YF **relative** risk of YF occurrence in second administrative locations given start locations in neighbouring countries. (a) where red indicates high risk and white/yellow indicates no/low risk. The relationship between the number of times a country appears in simulations, and the median risk score standardised by the number of second administrative locations from start locations in neighbouring countries. (b). Only median risk greater than 0.01 is displayed on the map.

### Underlying outbreak risk in the presence of a sylvatic reservoir

In addition to the scenario considered in the main text where YF virus spreads following the introduction of a single infection, based on estimate R_0_ values (section 2.3), we consider the speculative scenario where an established YFV sylvatic reservoir is present in the countries considered, by running the model with all 1000 sets of values of both R_0_ and *λ*_*S*_ estimated from environmental covariates (median values shown in Fig. 2 and again recording the proportion of instances in which an outbreak occurs in the target year. The results are shown in Fig. 9. In the presence of a sylvatic reservoir, the outbreak risk becomes very high (¿99%) across most regions. This may be considered a worst case scenario where YFV becomes endemic in the considered countries.

**Fig 9.**
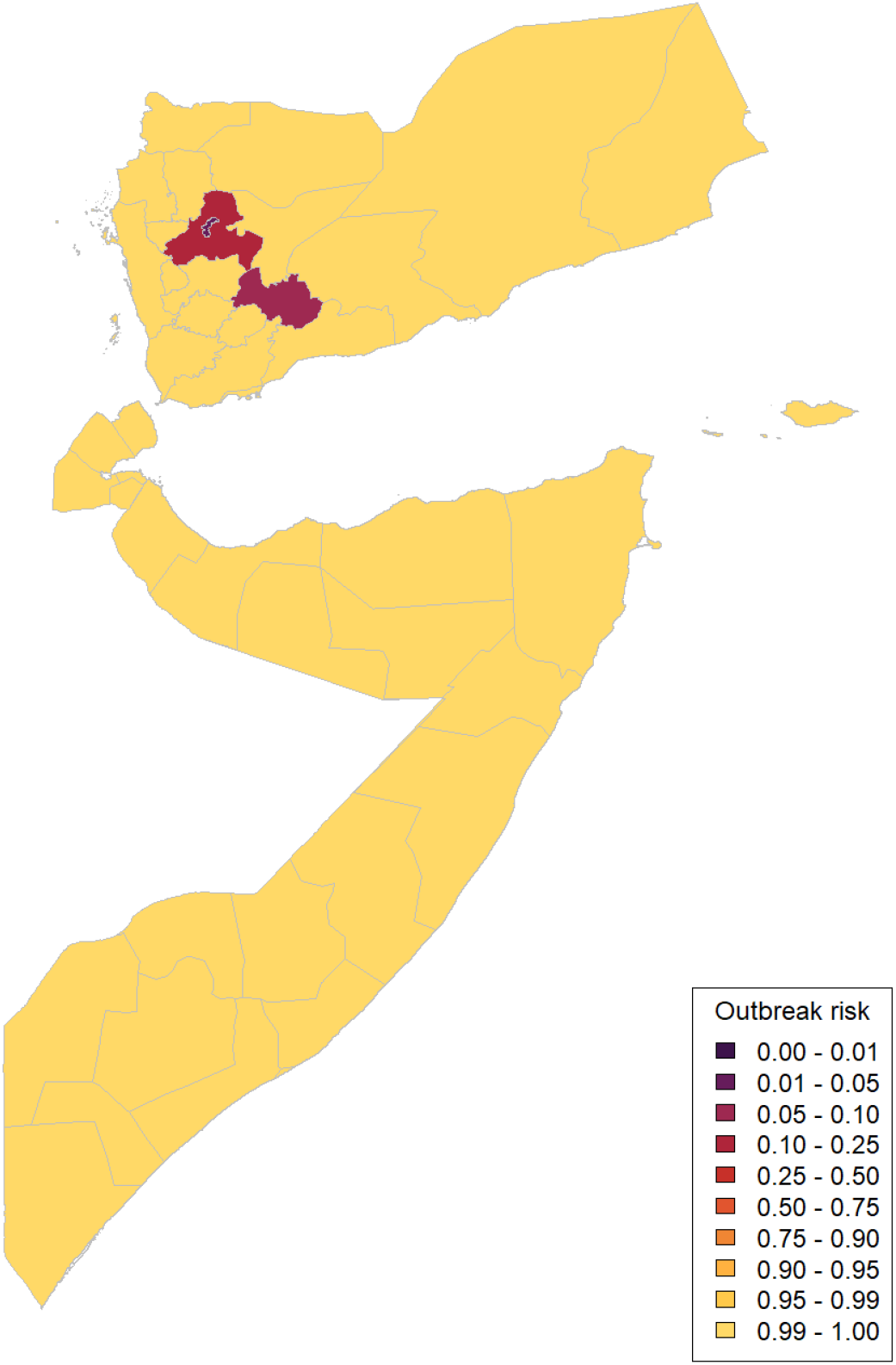
Maps of outbreak risk in first-level subnational administrative regions in Djibouti, Somalia and Yemen in 2023 as calculated by testing whether an outbreak propagates given the underlying estimates of sylvatic spillover force of infection and basic reproduction number from [4].

### Relative outbreak risk - alternate

**Fig 10.**
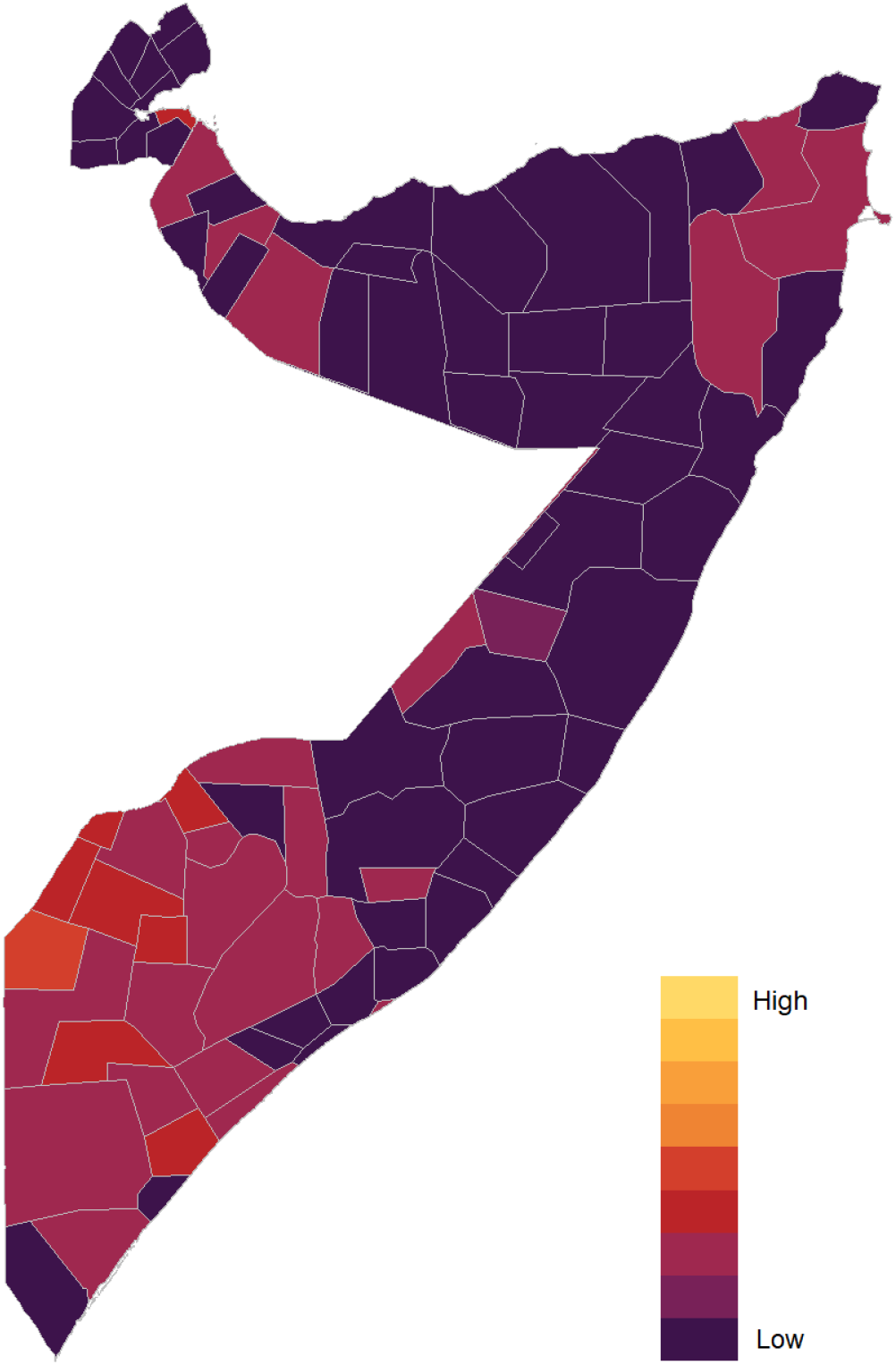
Map of relative outbreak risk in second level subnational administrative regions in Djibouti and Somalia in 2023 calculated via multiplying median introduction risk scores for each region by the underlying outbreak risk based on a single introduced case for the corresponding first level subnational administrative region

